# Benchmarking homologous recombination deficiency algorithms for prediction of clinical outcome in ovarian cancer

**DOI:** 10.1101/2025.10.08.25337487

**Authors:** Lasse Ringsted Mark, Inge Søkilde Pedersen, Anne Krejbjerg Motavaf, Henrik Bygum Krarup, Karen Dybkær, Ida Burchardt Egendal, Helle D Zacho, Mads Thomassen, Martin Bøgsted

**Author notes:** Corresponding author: Lasse Ringsted Mark.

## Abstract

Homologous recombination deficiency (HRD) is a predictive biomarker for response to platinum-based chemotherapy in ovarian cancer. In this retrospective study, we benchmarked three HRD detection algorithms - CHORD, ShallowHRD, and OvaHRDscar - alongside BRCA1/2 mutation status in a cohort of 100 patients with high-grade serous ovarian carcinoma (HGSC). HRD status was derived from whole genome sequencing of tumor/normal samples, and progression-free survival (PFS) was used as the primary endpoint.

All three HRD algorithms showed a statistically significant association with improved PFS. In multivariate Cox regression models adjusted for age, FIGO stage, tissue type, and neoadjuvant chemotherapy, HRD-positive status was significant associated with reduced hazard of progression or death: OvaHRDscar (HR = 0.41, 95% CI: 0.25–0.67), ShallowHRD (HR = 0.50, 95% CI: 0.31–0.79), and CHORD (HR = 0.47, 95% CI: 0.24–0.94). In contrast, BRCA1/2 mutation status did not show a significant association (HR = 0.64, 95% CI: 0.35–1.16).

These findings support that HRD algorithms may aid in the diagnostic assessment of HRD and support broader genomic profiling to enhance clinical decision-making. Future studies should focus on refining algorithm thresholds and validating these results in larger, multi-center cohorts to facilitate clinical translation.

## Introduction

Homologous recombination deficiency (HRD) is a defect in the homologous recombination repair (HRR) pathway, which is a critical repair mechanism for accurate repair of DNA double stranded breaks (1,2). Several types of cancers have been strongly associated with HRD especially ovarian and breast cancer, which are also known to have frequent alterations in HRR related genes, such as BRCA1 and BRCA2. Other cancers associated with HRD are, but not limited to, endometrial, pancreatic, and prostate cancer (1). Clinical trials have shown that patients harboring pathogenic BRCA variants or having a high genomic instability score (GIS), which are common markers for HRD, experience good response from platinum-based chemotherapies and Poly (ADP-ribose) polymerase inhibitors (PARPi) (3–5).

However, today there is no universal and reliable biomarker to detect HRD. Historically, pathogenic BRCA variants have been the go-to standard of assessing HRD, and more recent studies have shown that HRD status can be assessed through a variety of assays and algorithms, including gene expression analysis, single nucleotide polymorphism (SNP), genomic aberration, functional assays, and algorithms based on a variety of such biomarkers (6). Furthermore, these methods each have their own advantages and limitations with respect to both clinical applications and patient selection. Comparative data on their predictive performance in a real-world setting remain limited (6).

In this study we aim to evaluate three different HRD detection algorithms in a retrospective cohort of ovarian cancer patients. By evaluating algorithms with respect to progression-free survival (PFS) in patients treated with platinum-based chemotherapies, we aim to shed light on the clinical utility of HRD testing both for guiding personalized treatment and for understanding variability among HRD algorithms.

## Methods

### Study Cohort

The study cohort consists of 100 patients from Aalborg University hospital, Denmark, diagnosed with high-grade serous carcinoma (HGSC) ovarian cancer between 2016 and 2021. Patients were original enrolled in routine clinical diagnostic of ovarian cancer and screened for pathogenic BRCA 1/2 variants to guide further treatment and assess potential inherited cancer risk. Oncological treatment and response information were extracted from the electronic patient journals and verified by an experienced oncologist from the Department of Oncology, Aalborg University Hospital, Denmark. Pathology information regarding FIGO stage and histological subtype were extracted from The Danish Pathology Data Bank by a pathologist from Department of Pathology, Aalborg University Hospital, Denmark. All information were merged with data from the Danish Gynecological Cancer Database (DGCD) database from the Danish Healthcare Quality Institute. DGCD is a multidisciplinary clinical quality database that receives reports from nurses, gynecologists, pathologists, and oncologists and includes patients who had been diagnosed with, among others, ovarian cancer after January 1, 2005, in Denmark. Data include surgical information, performance indicators, and anthropometric and biochemistry measures. DNA from tumor and matched normal blood samples were obtained from a diagnostic biobank at Department of Molecular Diagnostics, Aalborg University Hospital, Denmark.

### Laboratory workflow

DNA was extracted from individually paired blood and tumor samples using MaxWell 16 FFPE Plus LEV DNA Purification kit (Promega) for archival tumor tissue (FFPE tissue), QIAamp Fast DNA Tissue Kit (Qiagen) for fresh frozen tumor tissue and QIAsymphony DNA Midi Kit (Qiagen) for EDTA blod. Extracted DNA was fragmented with Covaris ME220 (PerkinElmer) to an average size of 200 base pairs. For library preparation SureSelect XT HS2 DNA Reagent kit from Agilent Technologies was used. Adapters and barcodes were attached followed by 3 cycles of polymerase chain reaction (PCR) amplification. The library was sequenced on an Illumina NovaSeq 6000. In this study the target sequencing depth were 60X for tumour and 30X for blood samples.

### Pipeline processing, variant calling

All samples were bioinformatically processed through nf-core/Sarek version 3.2, a community curated pipeline within the nf-core framework, designed for pre-processing, germline and somatic variant calling as well as functional annotation (7,8). Sarek was configured to call both germline and somatic variants using Strelka2 and estimate microsatellite instability (MSI) with MSIsensor-pro (9,10). Default settings were applied. Structural variants were called using control-FREEC, Manta, and ASCAT(11–13). Structural variant calling was not called as part of Sarek but implemented individually based on best practice defined by the individual tool.

### Variant annotation

The clinical implication of variants related to HRR were assessed. The list of genes was extracted from Reactomes Pathway Browser category *“HDR through Homologous Recombination”* (14) and comprises 69 genes. The list can be found in Supplementary Materials. Germline and Somatic variants were analysed using VarSeq™ v2.6.2 (Golden Helix, Inc., Bozeman, MT, www.goldenhelix.com) and classified with the build-in annotation tool implementing the ACMG guidelines for variant interpretation and classification (15,16). A criterion for all germline variants were that they should be marked as PASS from the variant caller and classified as either likely pathogenic or pathogenic. These variants were further validated by a molecular biologist (ISP) from Department of Molecular Diagnostic, Aalborg University hospital, Denmark using gene-specific guidelines for variant classification where available within the Clinical Genome Resource (clinicalgenome.org) framework. For somatic variants the same criterions were applied with the addition that variants should be present in a minimum of five alignment reads and have an allele frequence of two percent or above.

### HRD testing

Three previously developed HRD algorithms, CHORD, ShallowHRD, and OvaHRDscar were implemented in this study(17–19). Each method categorized samples into HRD and non-HRD groups, with ShallowHRD including an additionally “borderline” category. CHORD is a random forest-based model which predict HRD based on mutational signatures, indels, and structural variants (19). In this study CHORD utilized somatic variants from Strelka2 and structural variants from Manta. ShallowHRD is based on copy number alterations(18) and was in this study implemented using control-FREEC as input. OvaHRDscar is a clinical outcome optimized reimplemented HRD algorithm estimating a genomic instability score, defined as the unweighted sum of the three structural variant types: large scale transitions (LST), Loss of heterozygosity (LOH), and Telomeric allelic imbalances (TAIs), which original were described by (Telli,2016) (5,17). In this study, OvaHRDscar scores were extracted from allele specific copy number output from ASCAT. All three algorithms output continuous scores or probabilities, hence a threshold is needed to categorize into HRD and non-HRD. In this study, we apply the thresholds from the original publications where each algorithm was developed. For OvaHRDscar a threshold of >54 is used to define HRD samples. For ShallowHRD two threshold are used. A score <15 is defined as non-HRD, a score between >= 15 and <=19 is defined as borderline, and a score >19 is defined as HRD. For CHORD a probability of >0.5 is defined as HRD(17–19).

### Statistical analysis

The primary endpoint of the study was PFS, defined as the time from the start of first-line treatment to disease progression, death, or loss to follow-up, whichever occurred first. Progression-free survival was illustrated using Kaplan-Meier curves. The Kaplan-Meier curves were estimated in python using the *KaplanMeierFitter* function from the package *lifelines* (20) Survival curves were compared by the Log-Rank test implemented by the function *logrank_test* in the same python package.

To estimate the effect sizes, a Cox proportional hazard model (Cox model) was fitted separately for each HRD algorithm and BRCA mutation status. The models were estimated in python using the function *CoxPHFitter* from the package *lifelines(20)*. To identify potential confounders a Directed Acyclic Graph (DAG) was constructed based on literature and domain knowledge. The Cox models were subsequently adjusted for potential confounders.

### Ethics

The study has been approved by the Danish National Committee on Health Research Ethics (Reference number: 2114546). The project has, according to the General Data Protection Rule (GDPR), been registered at the North Denmark Region’s list of projects handling person sensitive data (Reference number: F2022-016).

## Results

Cinical characteristics of the patients, including patient age at diagnosis, BMI, FIGO stage, tissue type, surgical outcome, and the use of neoadjuvant chemotherapy as well as molecular characteristics such as sequencing coverage, MSI status, and mutational status of germline and somatic BRCA1/2 and HRR genes can be seen in Table 1. Data are presented across the three HRD classification methods ShallowHRD, OvaHRDscar, and CHORD and HRD status based on BRCA mutational status. Of the 100 patients, 72 patients were diagnosed with FIGO stage III and 28 with FIGO stage IV. Tumor DNA were derived from either formalin-fixed paraffin-embedded (FFPE) tissue (n = 66) or fresh frozen (FF) tissue (n = 34). Of the 100 patients, 51 underwent macroradical resection, defined as surgical discretion at operation. Furthermore, 59 patients received neoadjuvant chemotherapy prior to surgical intervention, illustrating a large proportion of the cohort was treated with preoperative platinum-based chemotherapy. High-throughput sequencing was performed on tumor and individually matched blood samples of which tumor samples were sequenced with a mean depth of 65.10X (SD = 18.90). In comparison, blood-derived normal samples had a sequencing depth of 31.53X (SD = 5.59). Variant calling and annotation identified pathogenic or likely pathogenic variants according to ACMG guidelines in HRR genes including BRCA1 and BRCA2, (15). Pathogenic or likely pathogenic germline BRCA1/2 variants were identified in 11 patients and somatic BRCA1/2 pathogenic or likely pathogenic variants were identified in 5 patients. Beyond BRCA variants, 2 patients harbored germline variants in other HRR-related genes, and 1 patient exhibited a somatic HRR gene pathogenic variant. Due to various reasons, it was not possible to apply all HRD algorithms on all 100 samples. It was not possible to run OvaHRDscar on two samples, due to ASCAT failed to estimate ploidy and tumor purity. Hence, a total of 98 samples with OvaHRDscar scores were included. It was not possible to apply CHORD to one sample, due to Manta not being able to call CNVs, why 99 samples were included. It was possible to calculate ShallowHRD scores for all samples, whereas eight samples were classified as borderline, 50 samples classified as non-HRD, and 42 samples classified as HRD samples.

**Table 1:**
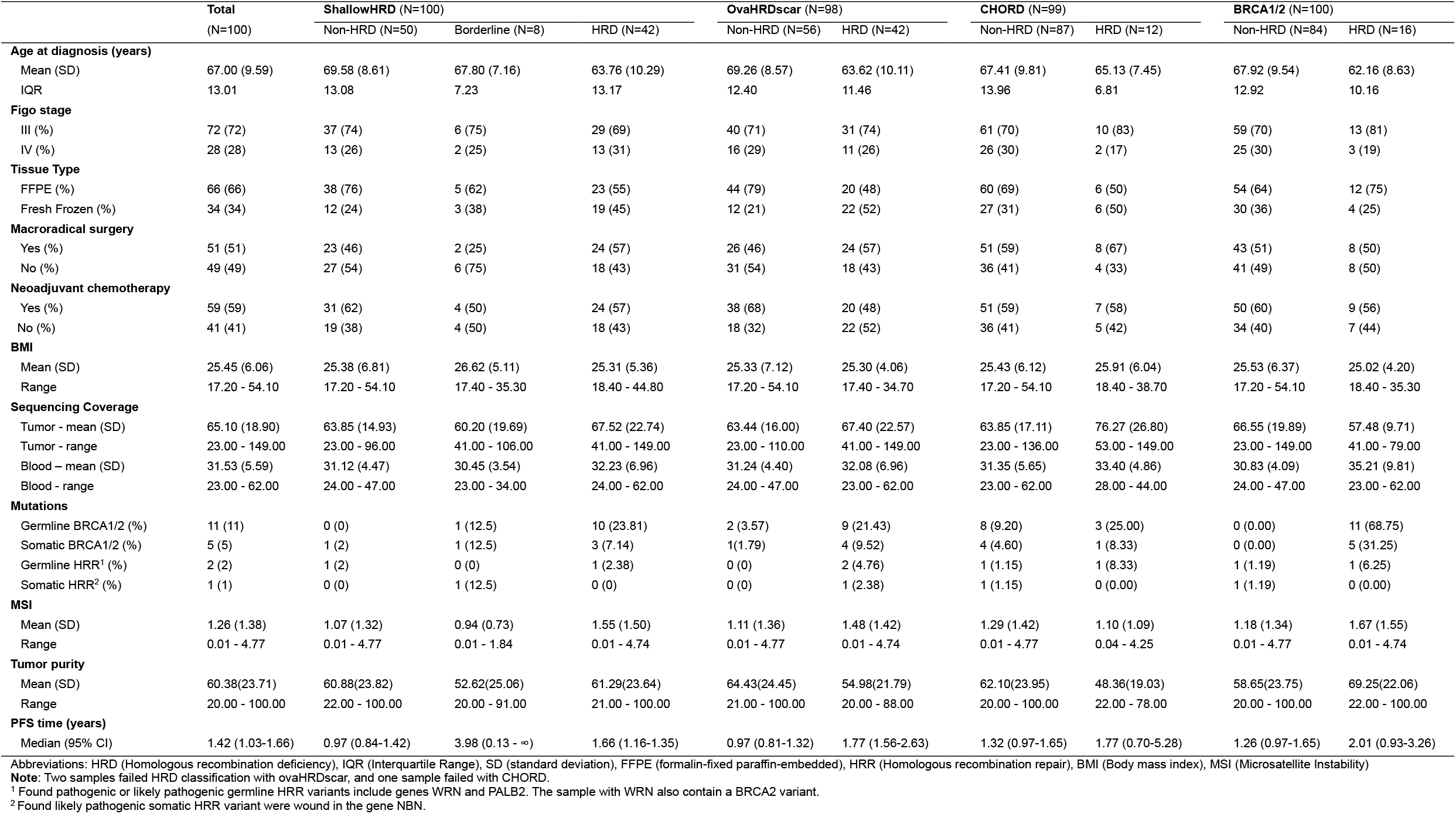
Clinical and molecular characteristics of the study cohort in general and across HRD classification methods.

To visualize and summarize which samples were classified as HRD samples and to assess common and unique samples a Venn diagram was constructed (Figure 1). The Venn diagram showed 42 samples were classified as HRD for ShallowHRD and OvaHRDscar, respectively, 12 samples for CHORD and 16 samples harboring somatic or germline BRCA variants. Notably, only 4 samples were classified as HRD for all three HRD algorithms and harboring BRCA variants. ShallowHRD and OvaHRDscar shared 32 samples classified as HRD. All 12 samples classified as HRD by CHORD were also classified as HRD by ShallowHRD. Interestingly, 8 and 9 samples were uniquely classified as HRD by ShallowHRD and OvaHRDscar, respectively. Two samples harbored BRCA variants, one being germline and one being somatic, without being classified as HRD by any HRD algorithm. The germline BRCA variant were however classified as borderline by ShallowHRD. Correlation diagrams between the individual scores can be found in Supplementary Figure 1

**Figure 1:**
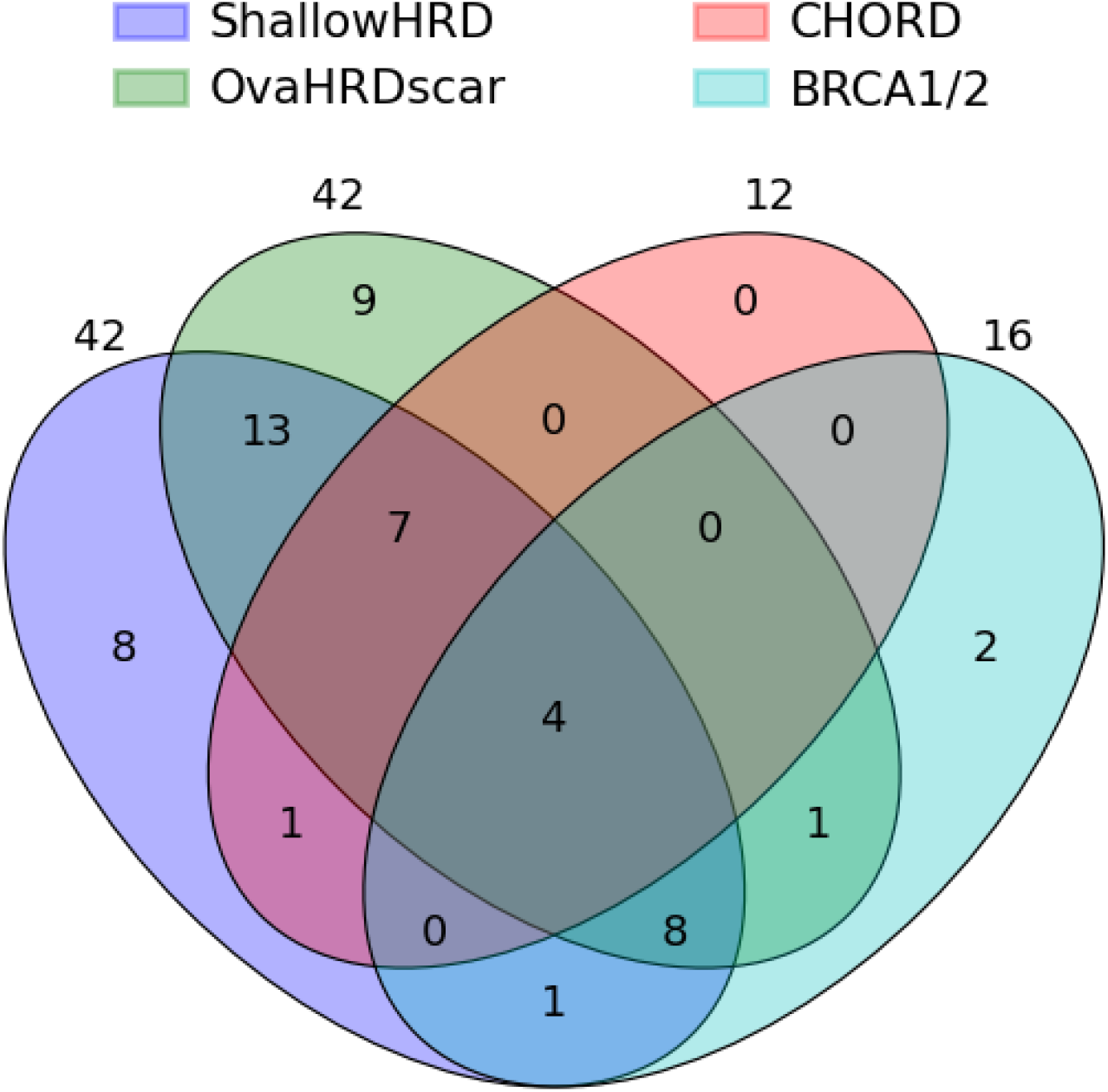
Venn diagram showing number of samples determined as HRD for each HRD algorithm and samples with pathogenic or likely pathogenic BRCA1/2 variants.

To evaluate the association between HRD classification and PFS, Kaplan-Meier curves are shown in Figure 2 for each HRD algorithm and for samples harboring BRCA variants. Samples classified as borderline were not included in the Kaplan Meier plot, due to the low numbers of samples. However, a Kaplan Meier plot including all three categories can be found in Supplementary Figure 2. A total of 16 samples harbored germline or somatic variants in either or both BRCA1 and BRCA2. The log-rank test comparing Kaplan-Meier curves revealed statistical significance in PFS between HRD-positive and HRD-negative groups for the three HRD algorithms CHORD, ShallowHRD, and OvaHRDscar, but not for BRCA1/2 mutational status.

**Figure 2:**
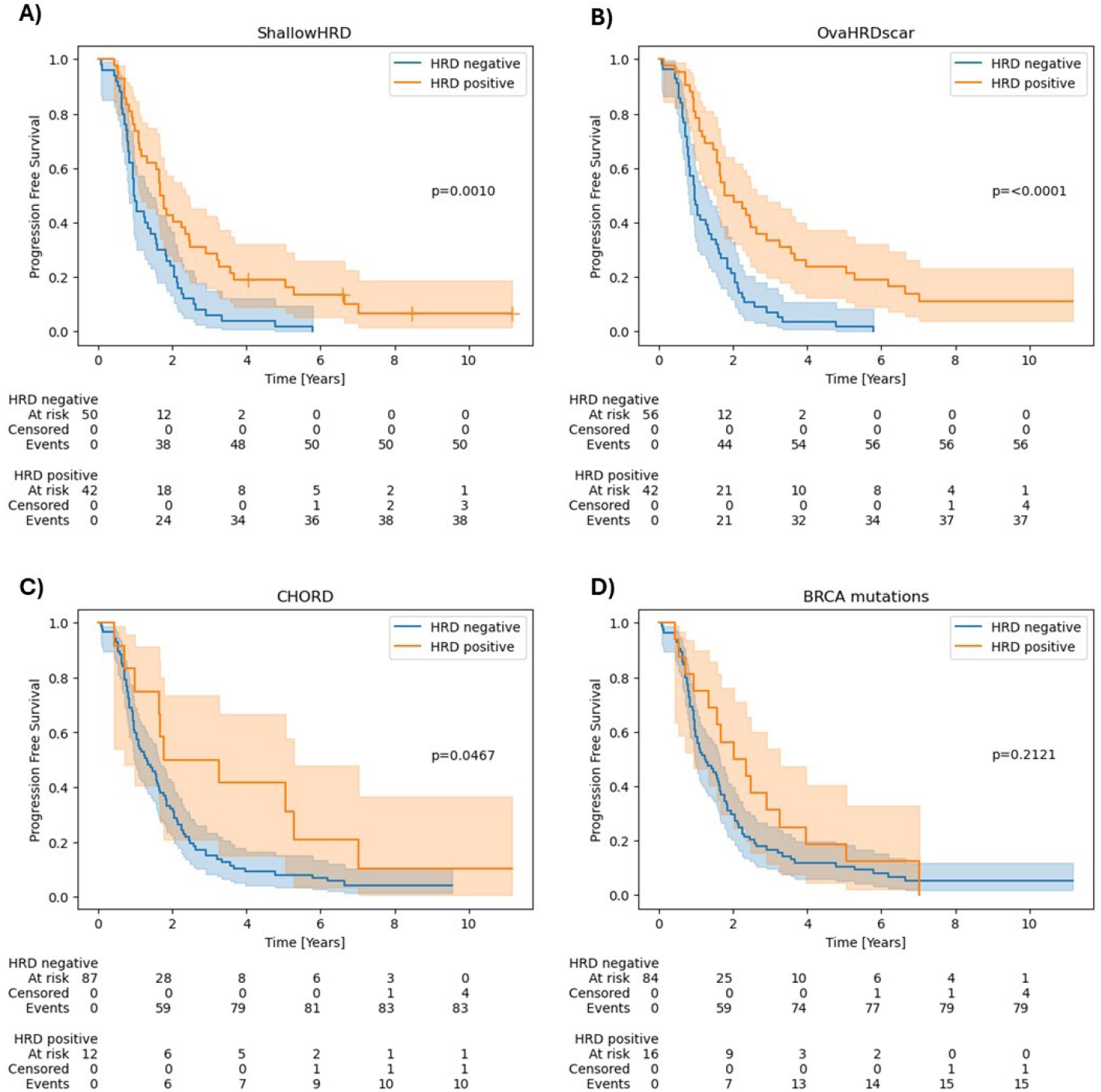
Progression free survival for A) ShallowHRD, B) OvaHRDscar, C) CHORD, and D) BRCA mutations. BRCA mutations include both somatic and germline pathogenic or likely pathogenic mutations.

In general, patients with HRD had longer PFS than non-HRD patients independent of HRD-algorithm. The median progression free survival time for ShallowHRD was 1.66 years (95% CI 1.16-2.35) for HRD-positive sample and 0.97 years (95% CI 0.84-1.42) for non-HRD samples. For OvaHRDscar the median progression free survival time was 1.77 years (95% CI 1.56-2.63) for HRD sample and 0.97 years (95% CI 0.81-1.32) for non-HRD samples. Median progression free survival time for CHORD was 1.77 years (95% CI 0.70-5.28) for HRD samples and 1.32 years (95% CI 0.97-1.65) for non-HRD samples. Lastly the median progression free survival time for BRCA mutated samples was 2.01 (95% CI 1.23-2.92) and 1.16 (95% CI 0.97-1.65) for samples without BRCA variants, (Table 1).

Boxplots of each HRD algorithm score with respect to tissue preservation medium, being either FFPE tissue or fresh frozen tissue, can be found in Figure 3. Furthermore, each category is divided by samples harboring BRCA variants and samples without BRCA variants. For OvaHRDscar, BRCA variant samples have a higher median score than BRCA wildtype samples. The median for the wildtype category is lower in FFPE than in fresh frozen tissue. Similar patterns are present for ShallowHRD. It is worth noting that there are very few samples with BRCA variants for fresh frozen tissue across all HRD algorithms. CHORD scores seem to follow a bimodal distribution. Scores are either low or high, with no or few samples scoring between 0.2 and 0.6. Furthermore, low scores are slightly higher for fresh frozen tissue than for FFPE tissue, especially for wildtype samples. High scores, however, seem to be similarly distributed.

**Figure 3:**
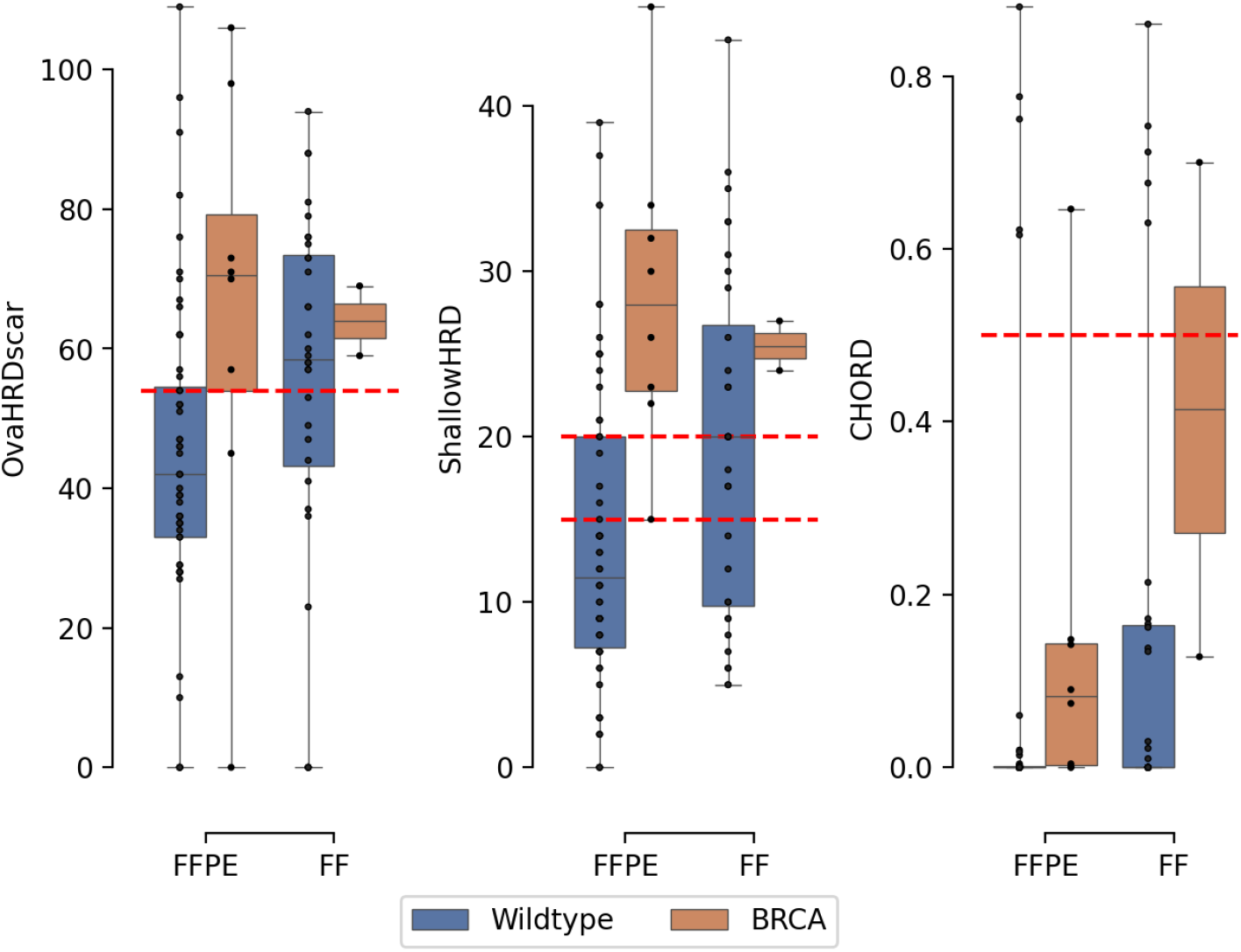
Boxplot showing the distribution of HRD scores across tissue type and mutational category. BRAC represent samples with pathogenic or likely pathogenic BRCA1 or BRCA2 mutations. Wildtype is samples with no known pathogenic or likely pathogenic BRCA1 and BRCA2 mutations. Red lines represent cut-off for the individual HRD algorithms. Abbreviations: FF (Fresh Frozen), FFPE (formalin-fixed paraffin-embedded)

We used a Cox model to estimate the effect sizes between HRD status and time to progression of first line treatment or death. For each HRD algorithm and BRCA mutational status, a multivariable cox model was estimated. Potential confounders were identified by construction of a DAG, Supplementary Figure 3. The Cox models were adjusted for identified confounders, being age at diagnosis, information on neoadjuvant chemotherapies, FIGO stage, and tissue type. Likewise, a univariate Cox model was estimated for each individual variable to assess the effect without adjusting for covariates in the multivariate model. There was a significant association between HRD status and event for OvaHRDscar both in the univariate model (HR of 0.38 CI: 95% 0.24-0.60) and the multivariate model (HR of 0.41 CI: 95% 0.25-0.67). The same was the case for ShallowHRD in both the univariate model (HR of 0.49 CI: 95% 0.32-0.76) and the multivariate model (HR of 0.50 CI: 95% 0.31-0.79). The Cox model for CHORD showed a non-significant Cox model for the univariate model (HR of 0.52 CI: 95% 0.27-1.00), and a significant multivariate model (HR of 0.47 CI: 95% 0.24-0.94. There was no significant association between HRD status and event for BRCA1/2 for neither the multivariable Cox model (HR of 0.64, CI: 95% 0.35-1.16) nor the univariate model (HR of 0.70, CI: 95% 0.40-1.22). The HR related to a positive HRD status was below 1 in both the univariate and multivariate model for all HRD algorithms and BRCA status and thereby indicating a decreased hazard or protective effect compared to patients with a negative HRD status. A summary of the estimated effect for the individual HRD algorithm and BRCA status as well can be found in Figure 4. The Cox model for the individual algorithms can be found in Supplementary Figure 4, Supplementary Figure 5, Supplementary Figure 6, and Supplementary Figure 7.

**Figure 4:**
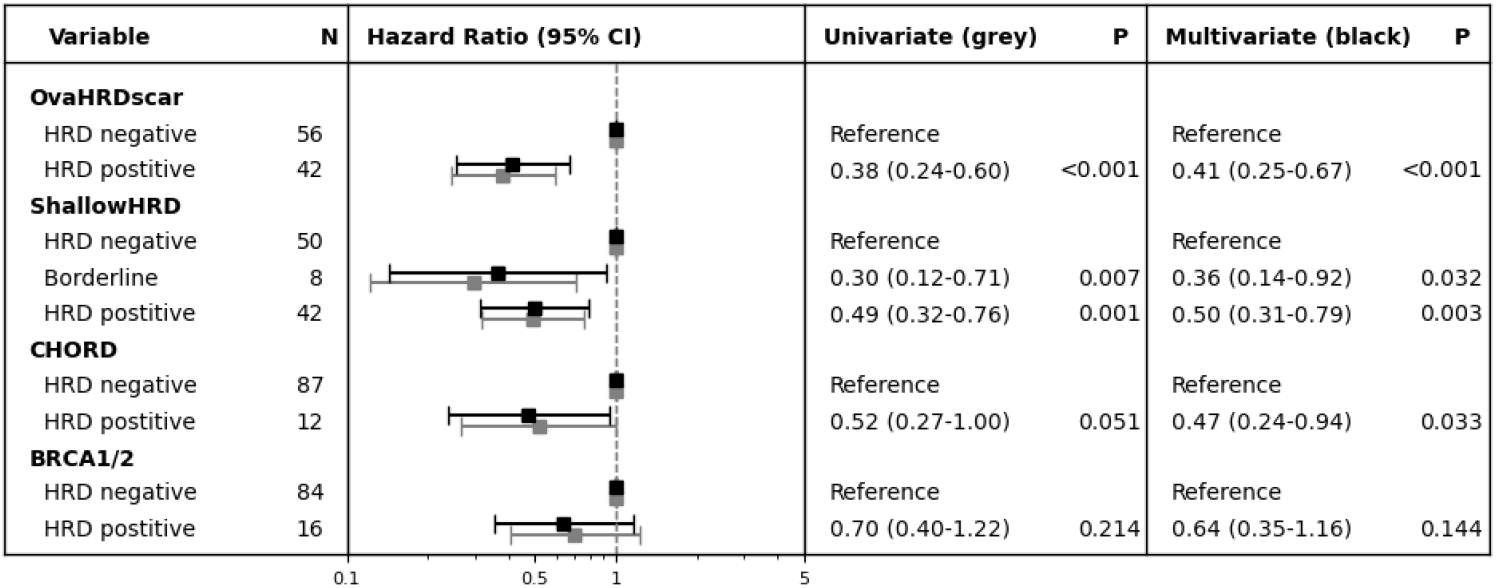
Summary of different Cox regression models estimated for the three different HRD algorithms and BRCA1/2 mutations. Hazard ratios based on a univariate (grey) and a multivariate (black) Cox models are shown with 95% confidence intervals and p-values. The univariate cox models have been estimated for HRD status only and the multivariate models include the variables: Tissue type, age, neoadjuvant chemotherapy, and FIGO stage.

Furthermore, we generated Kaplan-Meier curves to compare outcomes between patients with BRCA variants and those with non-BRCA variants, stratified by HRD status. This allowed us to assess whether HRD status impacts PFS differently depending on the presence or absence of BRCA mutations. Notably, in samples assessed using ShallowHRD and OvaHRDscar, HRD positive samples without BRCA variants showed a PFS similar to that observed in samples with BRCA variants, see Figure 5.

**Figure 5:**
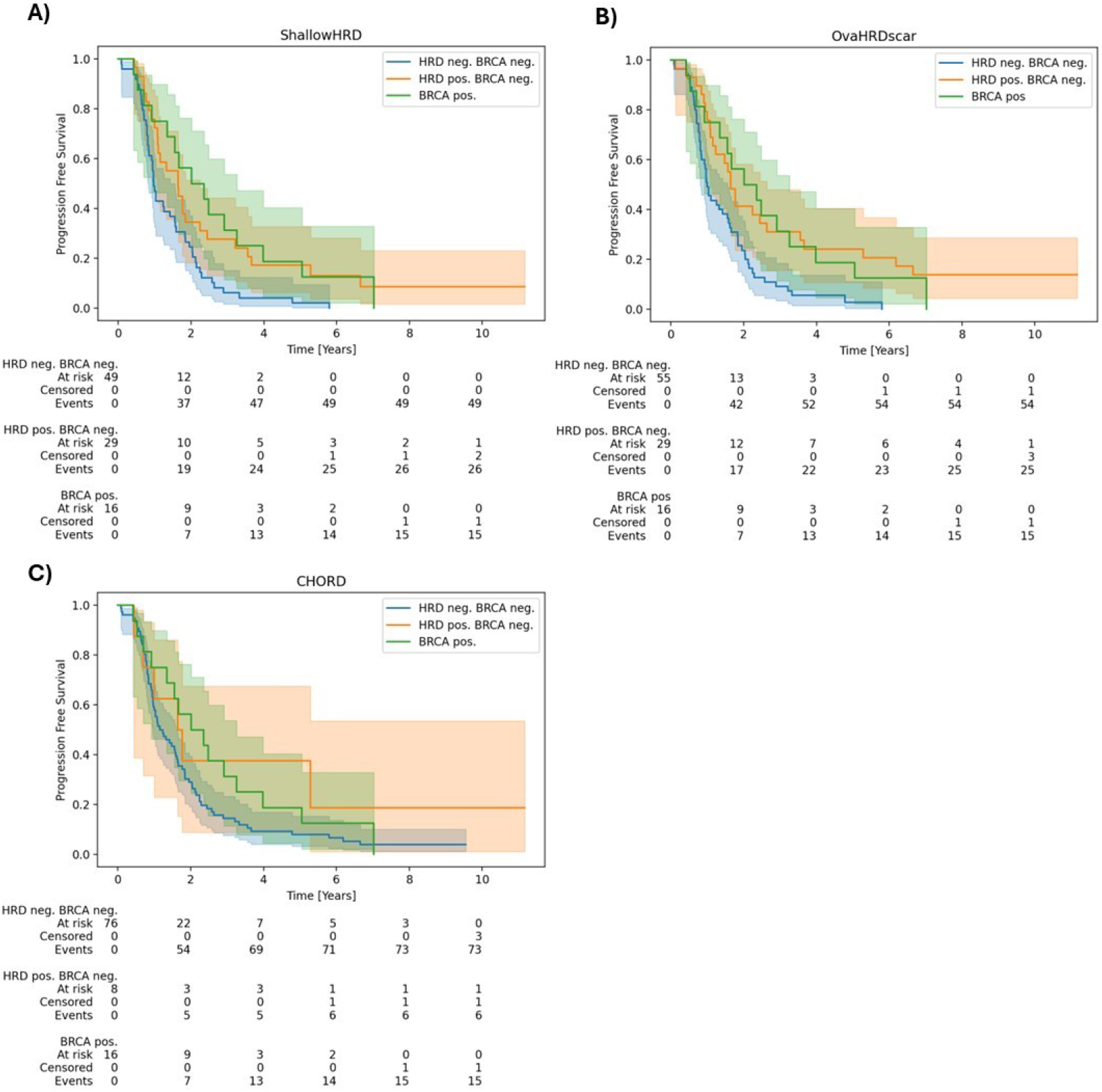
Progression free survival for A) ShallowHRD, B) OvaHRDscar, and C) CHORD, when considering BRCA mutations as a separate group, independent of HRD score/classification. BRCA mutations include both somatic and germline pathogenic or likely pathogenic mutations.

## Discussion

In this study, we evaluated the predictive utility of three HRD detection algorithms, namely CHORD, ShallowHRD, and OvaHRDscar, as well as BRCA1/2 mutation status, in relation to PFS following platinum-based chemotherapy in a retrospective cohort of 100 patients diagnosed with HGSC ovarian cancer. Even though the three HRD algorithms demonstrated varying degrees of performance, each showed a statistically significant association between HRD status and PFS defined as time from the start of first-line treatment to disease progression, death, or loss to follow-up. BRCA1/2 mutation status alone did not have significant association to PFS.

The comparison of the three different HRD algorithms showed both similarities and dissimilarities in classification of HRD samples. In general, there were a good concordance between OvaHRDscar and ShallowHRD, while most samples classified as HRD by CHORD, were also classified as HRD by ShallowHRD and OvaHRDscar. Not all BRCA-mutated tumors were classified as HRD samples by all algorithms, although BRCA1/2 variants in general are a well-established marker of HRD (1,21,22). However, both OvaHRDscar and ShallowHRD classified a considerable proportion (13/16) of BRCA mutated samples as HRD samples while also classifying additional 29 BRCA wildtype samples as HRD. This support the theory that HRD extends beyond BRCA variants and supports the use of broader genomic profiling to capture the full spectrum of HRD-related alterations, which has also been discussed and underlined in several other studies (1,6,21,22).

In the study which describes the development and optimization of OvaHRDscar, three cohorts were analyzed, revealing BRCA variants/deletion rates of 9%, 16%, and 22%, respectively. The corresponding proportions of HRD-positive cases were 56%, 50%, and 47%, (17). In the cohort analyzed in the present study, 16% of patients harbored somatic or germline BRCA variants, and 42% were classified as HRD positive based on both the OvaHRDscar and ShallowHRD algorithms. While the proportion of BRCA variants in the present study cohort is comparable to the proportions reported in another study, the proportion of HRD-positive cases was slightly lower(17). One explanation for this discrepancy could be the type of tissue preservation, as we observed slightly higher OvaHRDscar and ShallowHRD scores for fresh frozen samples compared to FFPE samples. With a general cut-off this will consequently influence the proportion of HRD positive classified samples. CHORD classified 6% of samples in a pan-cancer cohort as HRD positive, of which 94% harbored pathogenic or likely pathogenic variants in minimum one of the genes BRCA1/2, PALB2, RAD51C (19). In this study ∼12% of samples were classified as HRD positive by CHORD of which 4/16 (25%) samples harbored BRCA1/2 variants, which are quite different proportions. A study has found that up to ∼40-50% of ovarian cancers are HRD samples (23). Therefore, CHORD appears to classify significantly fewer samples as HRD, whereas OvaHRDscar and ShallowHRD perform at a comparable level. Another interesting thing is that samples classified as HRD by ShallowHRD, OvaHRDscar, and BRCA1/2 all had higher mean MSI values compared to the corresponding non-HRD samples. This was not the case for CHORD, where MSI values in general were lower in the non-HRD group compared to the HRD group. This is interesting as MSI samples previously have been a challenge to classify correctly by CHORD, (19). ShallowHRD did as the only algorithm include a borderline category, which also showed a significant association with PFS and similar hazard ratio as samples classified as HRD, indicating that cutoff thresholds could be optimized when evaluated in relation to PFS. This might also have been addressed in the updated version, *ShallowHRDv2*, which has been evaluated in a phase-III clinical trial, but unfortunately code and software are not publicly available at present, (24).

Both multivariate and univariate Cox models were fitted for each HRD algorithm and BRCA variants. All three multivariate Cox models for the individual HRD algorithms showed significant association between HRD score and PFS, whereas the BRCA Cox model did not. OvaHRDscar has previously shown significant association to PFS in patients treated with platinum-based chemotherapies, (17) Studies have shown a mixed association between BRCA mutational status and either PFS or platinum-based chemotherapies response. Two studies found a significant association (25,26) another study did not demonstrate any significant association(17). In accordance with the latter study, our findings showed a non-significant association between BRCA mutated samples and PFS. This might be due to low power caused by the small number of samples harboring BRCA variants.

Samples classified as HRD-positive by both ShallowHRD and OvaHRDscar exhibited a PFS nearly comparable to that of patients with BRCA variants, Figure 5. This finding shows that HRD status, independent of BRCA mutation status, is a strong predictor of response to platin chemotherapies, which has also been found in other studies, (27,28). Given that previous studies have shown a correlation between response to platinum chemotherapies and therapies targeting DNA damage repair in general, it is plausible that these findings may extend to a wider range of treatments, (28).

A key strength of this study is the comprehensive integration of clinical, pathological, and genomic data resulting in a well characterized patient cohort of HGSC ovarian cancer patients. The use of a public available community curated bioinformatics pipeline for primary and secondary analysis further enhances the reliability and reproducibility of results. Although conducted as a retrospective, single-center study, these findings provide a strong foundation for future, multi-center validation studies to confirm generalizability. While the application of previously established thresholds may require further optimization for our specific cohort or application, potentially influencing classification accuracy, the findings of this study remain significant. The variability in performance between HRD algorithms emphasizes the importance of algorithm selection to maximize clinical utility. In addition to further validation, standardizing HRD assessment remains essential for transferring these methods into clinic. Future studies should aim to validate these findings in larger multi-center cohorts to enhance generalizability. Additionally, refining thresholds for individual HRD algorithms is essential for various applications, including clinical implementation. Finally, a continued inclusion of emerging HRD algorithms is crucial to ensure that HRD assessment remains optimized, accurate, and clinically relevant.

## Conclusion

The study showed that the HRD detection algorithms, CHORD, ShallowHRD, and OvaHRDscar, are significantly associated with PFS in HGSC patients treated with platinum-based chemotherapy. In contrast, BRCA1/2 mutational status did not show a significant association, possibly due to the relatively few samples harboring BRCA1/2 variants. This supports that the incorporation of broader genomic profiling may enhance the overall HRD assessment. This study supports that HRD as a biomarker have great potential in stratifying patients, while also highlights the importance of selecting the appropriate HRD algorithm for clinical use. The substantial difference in performance among HRD algorithms underline the importance of algorithm selection, while also emphasizing the need to consider the tissue preservation medium. Future research should aim to refine algorithm thresholds based on PFS and further investigate the impact of tissue preservation methods. Continued evaluation of emerging HRD algorithms is always essential to ensure optimized and clinically relevant HRD assessment.

## Data Availability Statement

Data generated during the current study are not publicly available, as they contain information that could compromise the privacy of the research participants. However, they are available from the corresponding author upon reasonable request and following review and approval by the Danish Health Research Ethics Committee.

## Author Approval

All authors have read and agreed to the published version of the manuscript.

## Supplementary Materials

List of HRR genes

RMI1, EME2, CHEK1, RAD51B, RAD51C, XRCC3, XRCC2, RAD1, HUS1, NBN, RAD51D, RAD17, UBB, UBC, PCNA, RPA2, RPA1, POLD1, RPA3, RFC4, RFC2, RFC1, BRCA1, RFC5, RFC3, POLD2, MRE11, DNA2, BRCA2, BLM, POLE2, SEM1, RPS27A, UBA52, RAD51, POLE, ATM, TOP3A, ATR, SPIDR, WRN, POLD3, GEN1, FIGNL1, RAD9B, PALB2, SLX4, ATRIP, TOPBP1, RAD50, KAT5, EME1, RAD51AP1, RMI2, MUS81, RAD9A, RBBP8, BARD1, SLX1A, RHNO1, BRIP1, POLD4, POLE4, POLE3, FIRRM, RTEL1, POLK, EXO1, POLH

**Supplementary Figure 1:**
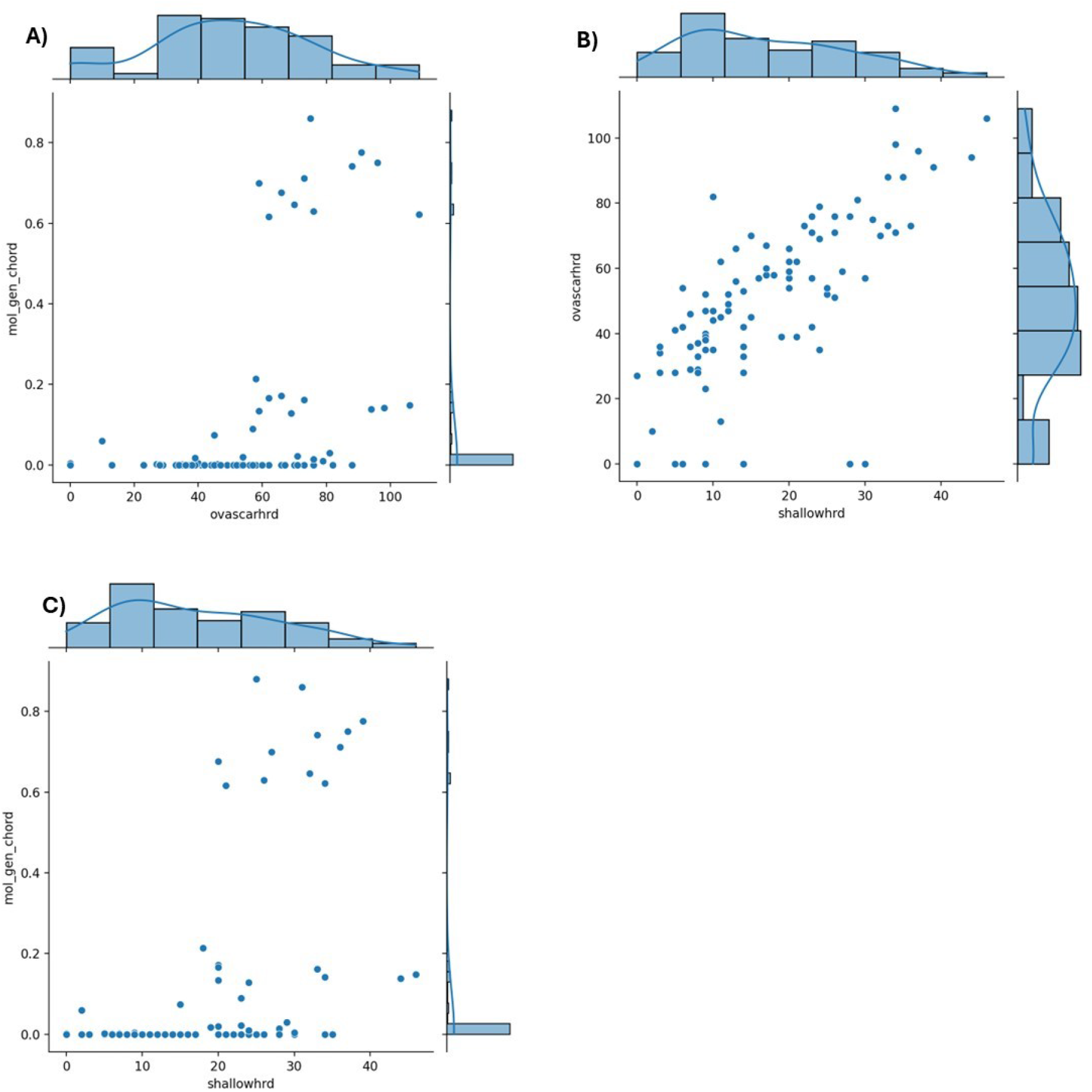
Correlation and distribution diagrams. A) Correlation between CHORD and ovaHRDscar scores. B) Correlation between ovaHRDscar and ShallowHRD scores. C) Correlation between CHORD and ShallowHRD scores.

**Supplementary Figure 2:**
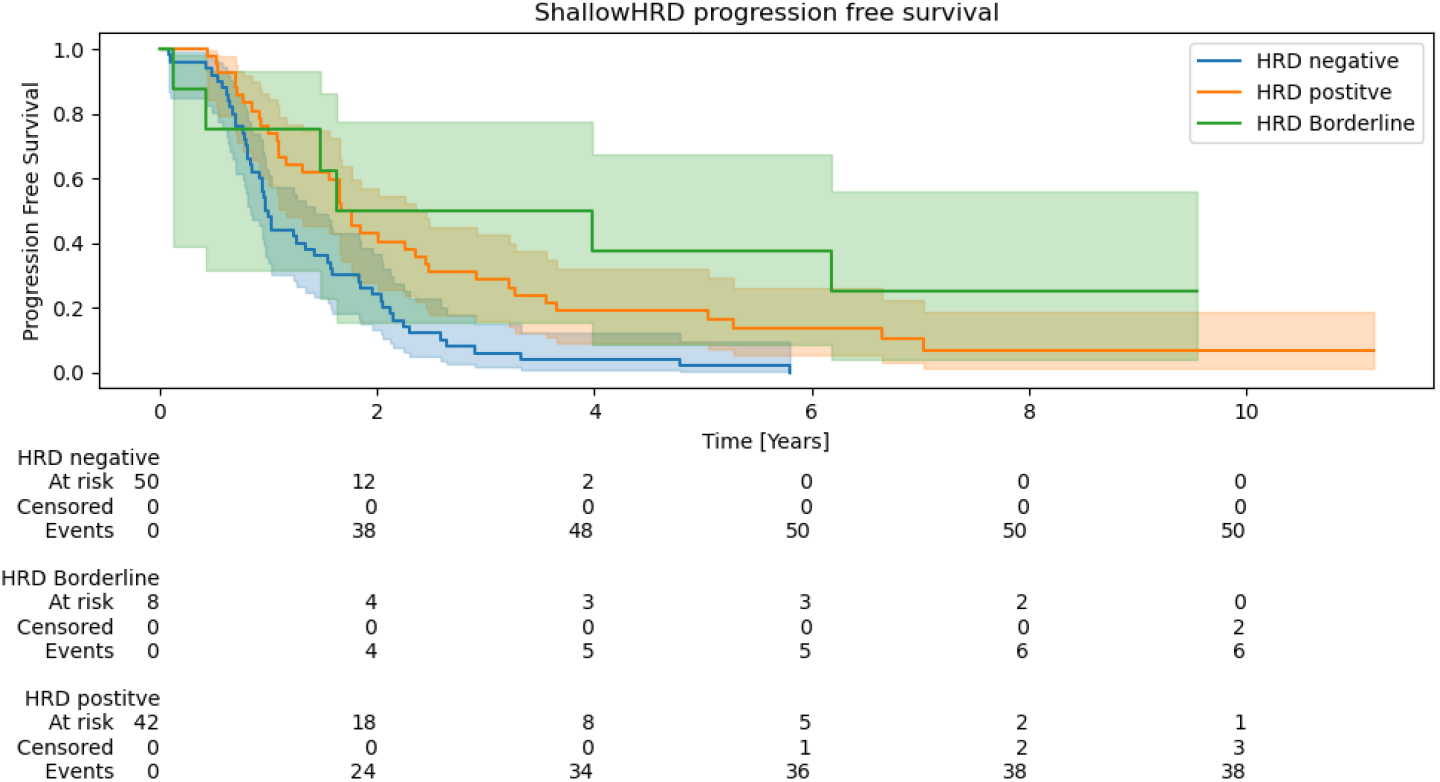
Progression free survival for ShallowHRD, including the three groups: HRD positive, HRD negative, and borderline.

**Supplementary Figure 3:**
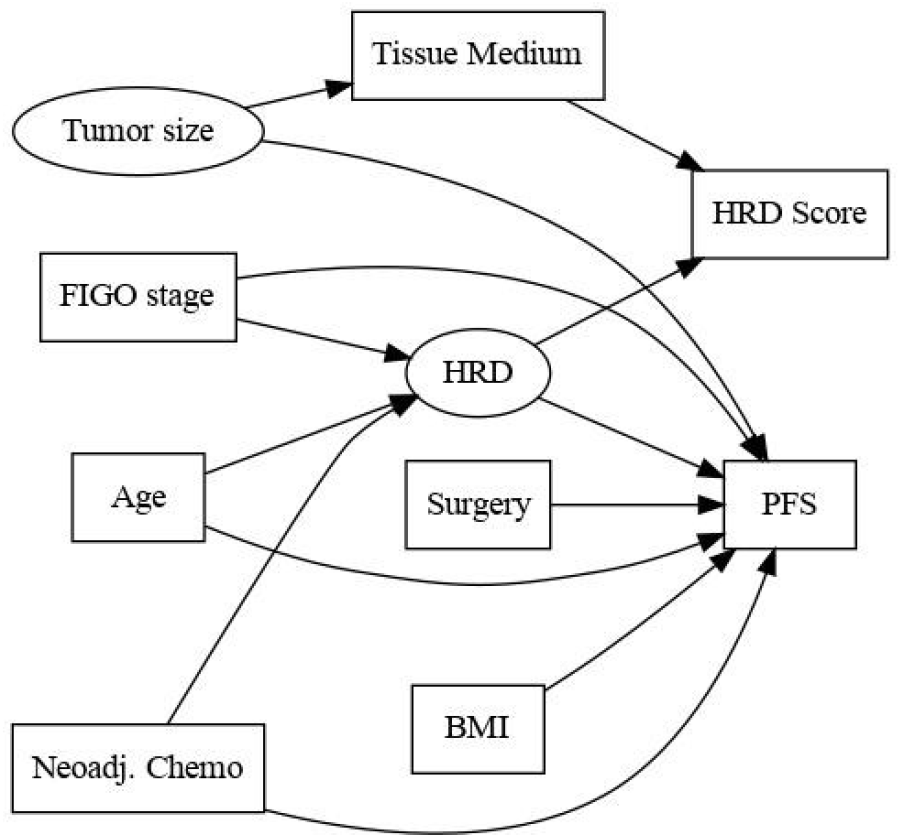
Directed Acyclic Graph (DAG) representing the hypothesized causal relationships among the variables in our study. Each node corresponds to a variable, and directed edges indicate assumed causal relationship. Rectangular shaped nodes represent observed variables and ellipses represent unobserved variables. This DAG are based on literature and domain knowledge and help identifying potential confounders.

**Supplementary Figure 4:**
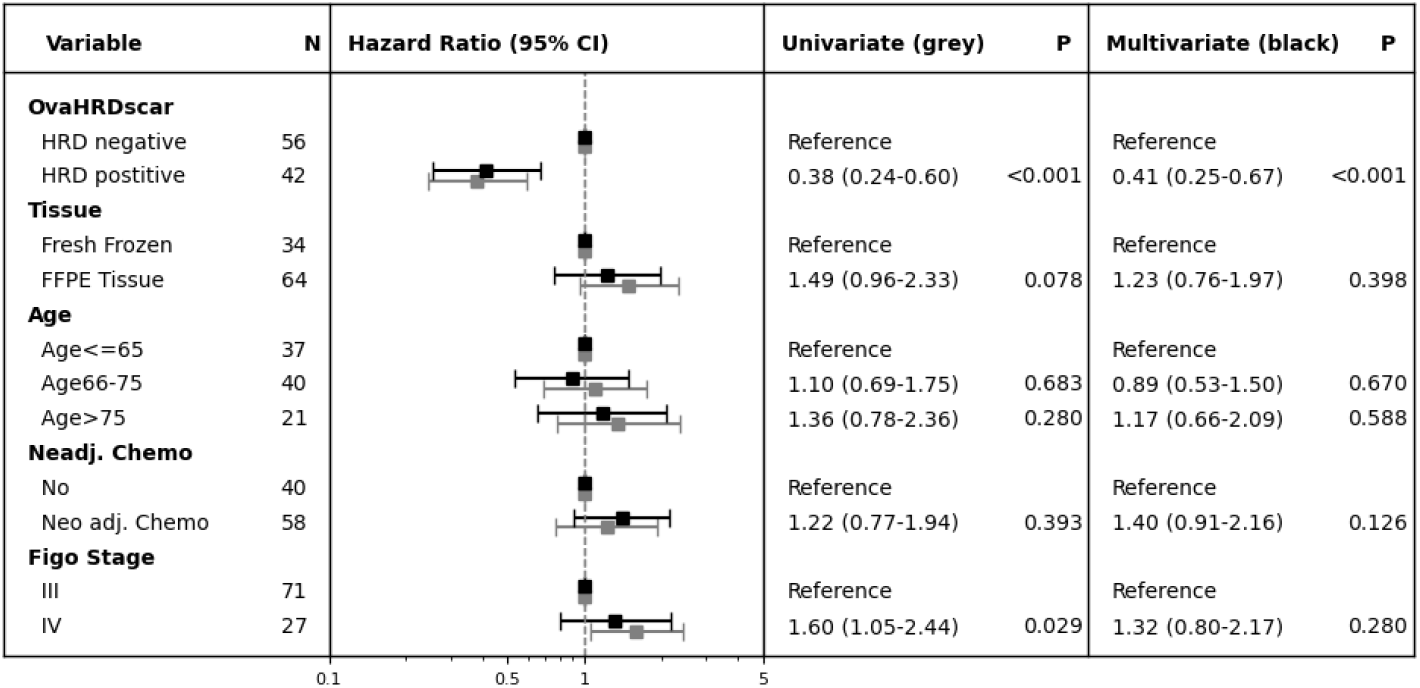
Cox regression model including the HRD algorithm OvaHRDscar and modelling the risk of disease progression in first line treatment or death. Hazard ratios based on a univariate (grey) and a multivariate (black) Cox regression model are shown with 95% confidence intervals and p-values. The univariate cox models been estimated for each variable.

**Supplementary Figure 5:**
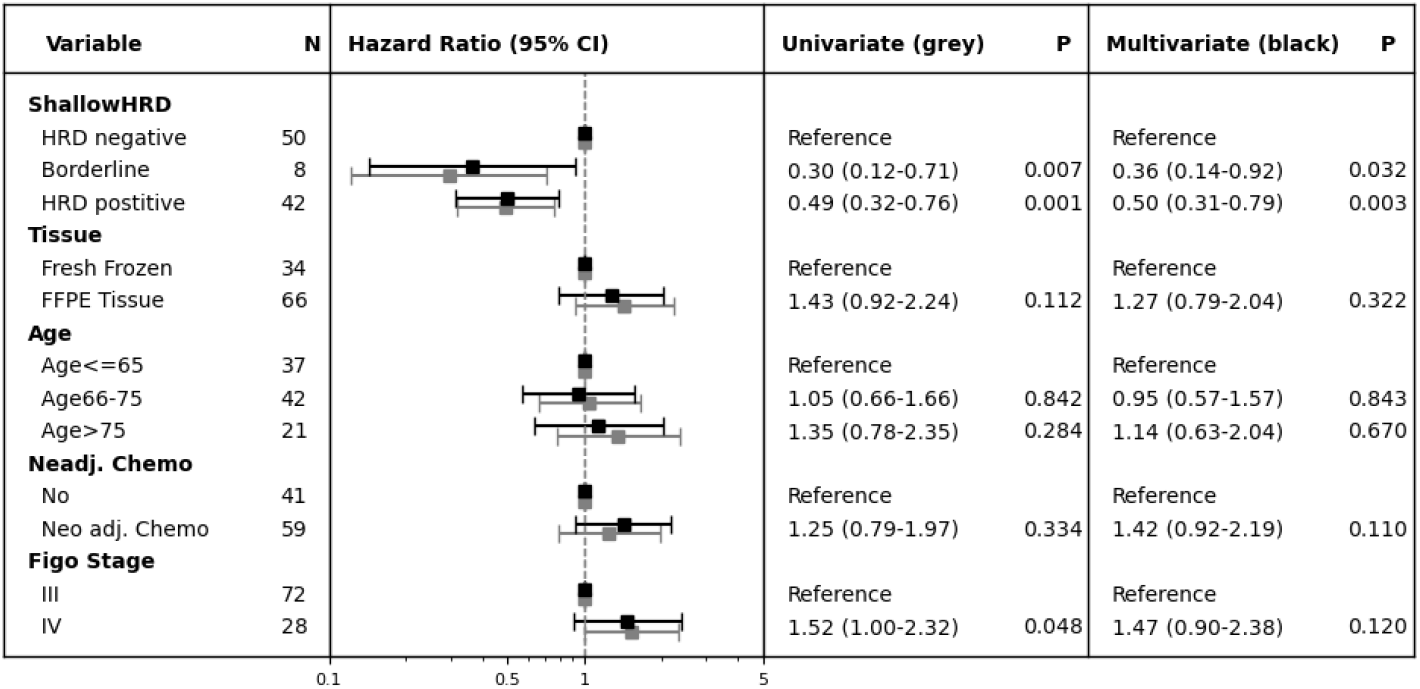
Cox regression model including the HRD algorithm ShallowHRD and modelling the risk of disease progression in first line treatment or death. Hazard ratios based on a univariate (grey) and a multivariate (black) Cox regression model are shown with 95% confidence intervals and p-values. The univariate cox models been estimated for each variable.

**Supplementary Figure 6:**
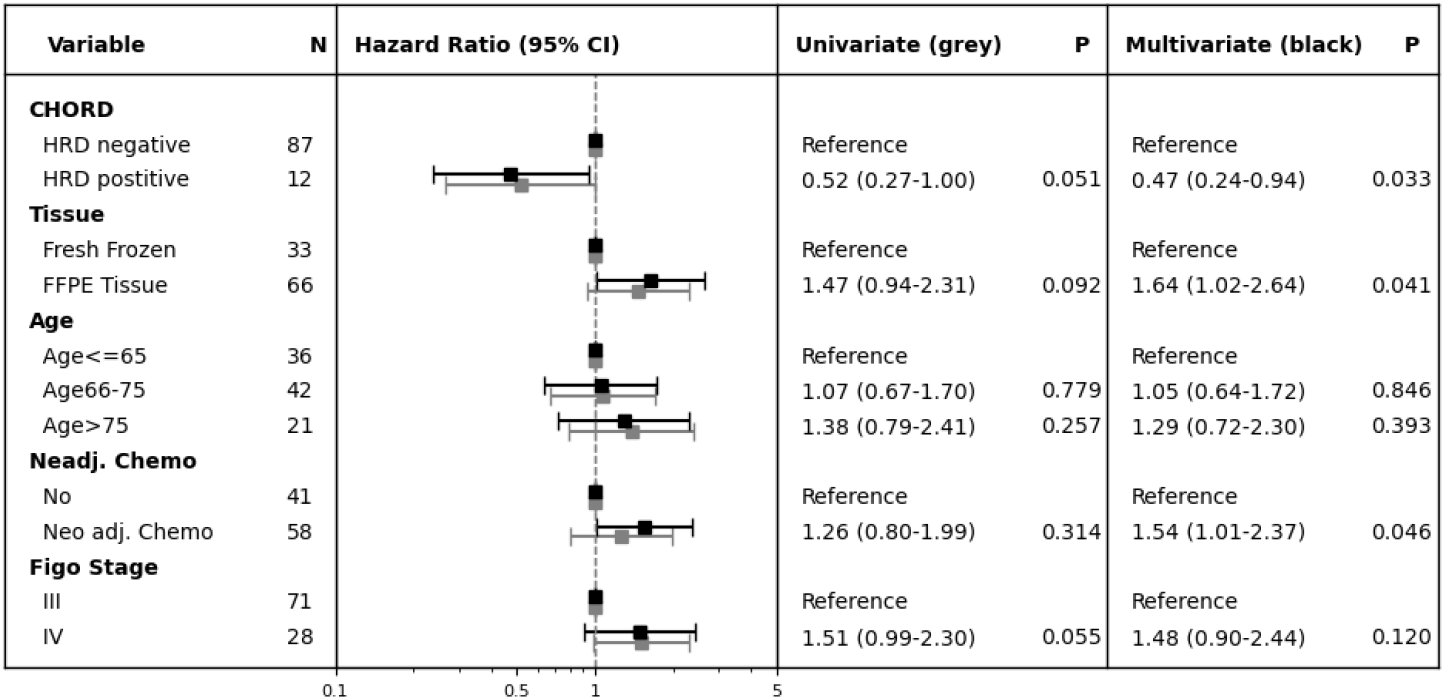
Cox regression model including the HRD algorithm CHORD and modelling the risk of disease progression in first line treatment or death. Hazard ratios based on a univariate (grey) and a multivariate (black) Cox regression model are shown with 95% confidence intervals and p-values. The univariate cox models been estimated for each variable.

**Supplementary Figure 7:**
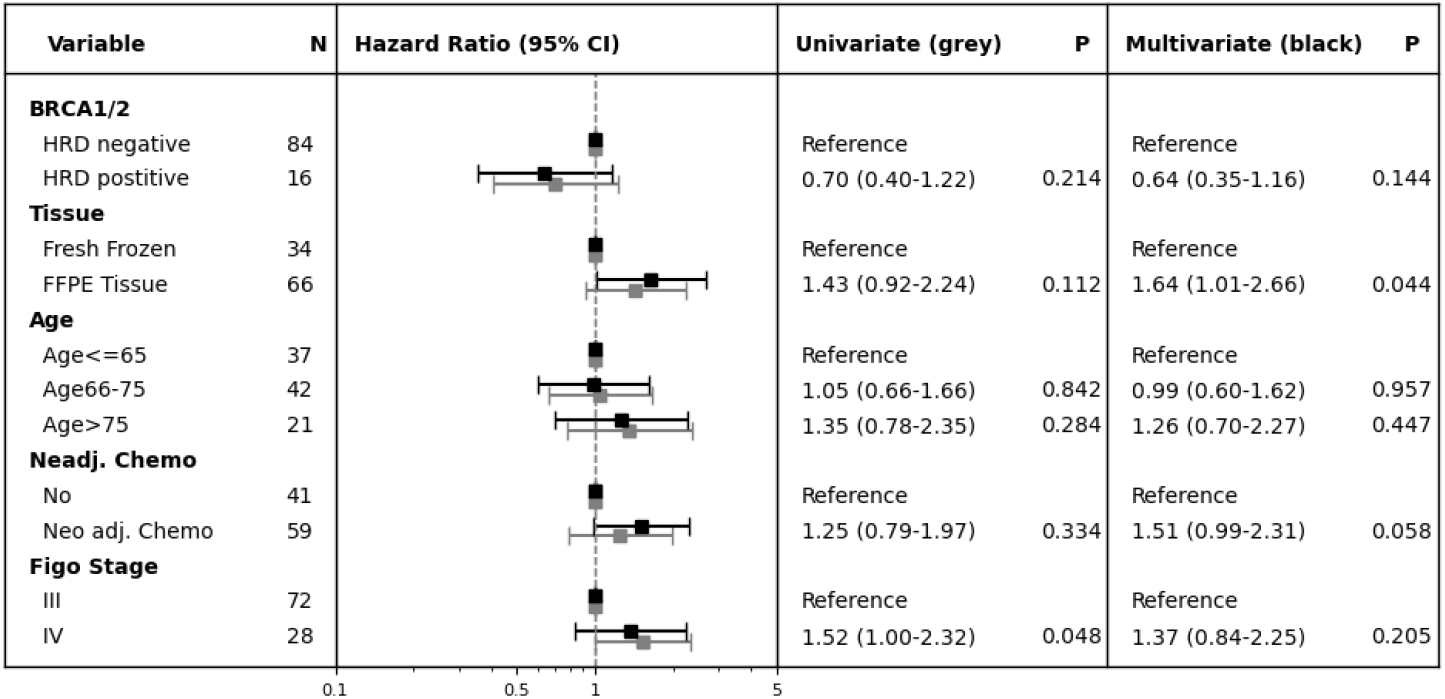
Cox regression model including BRCA mutation and modelling the risk of disease progression in first line treatment or death. Hazard ratios based on a univariate (grey) and a multivariate (black) Cox regression model are shown with 95% confidence intervals and p-values. The univariate cox models been estimated for each variable.

